# Dietary Diversity and Underweight Among Women Aged 15–49 Years in Cambodia: Analysis of the 2021–22 Cambodia Demographic and Health Survey

**DOI:** 10.64898/2026.10.13.26365381

**Authors:** Chantrea Sieng, Samnang Um, Sovandara Heng

## Abstract

**Aim:** This study aimed to estimate the prevalence and distribution of minimum dietary diversity for women (MDD-W) and underweight/thinness and examine sociodemographic and dietary factors associated with underweight/thinness among Cambodian women aged 15–49 years.

**Methods:** We conducted a cross-sectional analysis of 9,181 women aged 15–49 years (9,262.5 weighted) from the 2021–22 Cambodia Demographic and Health Survey. Survey-weighted descriptive statistics and provincial mapping were used to describe MDD-W, food-group consumption and underweight/thinness. Rao–Scott chi-square tests assessed group differences, and survey-weighted logistic regression estimated factors associated with underweight/thinness. MDD-W and household wealth were retained a priori in the adjusted model, while other covariates with an overall crude Wald P value <0.20 were included.

**Results:** The analysis included 9,181 women (9,262.5 weighted). Overall, 57.2% achieved MDD-W, and 6.7% had underweight/thinness; 22.8% were overweight, and 5.7% had obesity. Underweight/thinness was highest among women aged 20–24 years (17.7%). Achieving MDD-W was not associated with underweight/thinness in crude analysis (OR 0.93, 95% CI 0.74 to 1.17; P=0.545). In the adjusted analysis, the association remained imprecise (aOR 0.86, 95% CI 0.67 to 1.09; P=0.219). Provincial estimates showed geographic variation in both MDD-W and underweight/thinness.

**Conclusion:** More than half of Cambodian women achieved MDD-W, while underweight/thinness coexisted with overweight and obesity. MDD-W was not clearly associated with underweight/thinness. These findings support monitoring dietary diversity, anthropometric status, and socioeconomic inequalities together in women’s nutrition.

**WHAT IS ALREADY KNOWN ON THIS TOPIC:** Minimum dietary diversity for women is a population-level indicator of dietary diversity and a proxy for micronutrient adequacy among non-pregnant women aged 15–49 years. Cambodia is experiencing a nutrition transition in which undernutrition coexists with increasing overweight and obesity.

**WHAT THIS STUDY ADDS:** Using nationally representative 2021–22 Cambodia DHS data, this study describes MDD-W, individual food-group consumption, age-appropriate underweight/thinness, the full nutritional-status distribution, and provincial variation among women aged 15–49 years.

**HOW THIS STUDY MIGHT AFFECT RESEARCH, PRACTICE OR POLICY:** The results can support nutrition surveillance and help identify population groups and geographic areas requiring closer attention. Because MDD-W captures dietary diversity rather than total energy intake, it should be interpreted alongside anthropometry and other indicators of diet and nutritional status.

## INTRODUCTION

Malnutrition among women of reproductive age remains an important public health concern because nutritional status influences women’s health and can affect pregnancy and intergenerational health ^1 2^. At the same time, many low- and middle-income countries are experiencing a nutrition transition in which persistent undernutrition coexists with overweight and obesity ^1 3 4^. This double burden requires surveillance approaches that consider both anthropometric status and diet quality ^3 4^.

Dietary diversity is one dimension of diet quality. The Food and Agriculture Organization (FAO) defines Minimum Dietary Diversity for Women (MDD-W) as consumption of at least five of ten defined food groups during the previous 24 hours among women aged 15–49 years ^5–7^. MDD-W was developed for population-level assessment and is used as a proxy for micronutrient adequacy rather than as a direct measure of energy or protein intake ^6 8^. Validation research has shown that food-group diversity indicators are associated with micronutrient adequacy, while work including Cambodian women has also evaluated the validity of practical MDD-W data-collection methods against weighed food records ^8 9^.

Cambodia has experienced substantial demographic, socioeconomic and nutritional change. Analyses of Cambodian DHS data from 2000–2014 documented persistent inequalities in women’s nutrition alongside a changing distribution of underweight and excess weight ^10^. The 2021–22 CDHS provides the most recent nationally representative platform for examining women’s nutritional status and related characteristics ^11^. However, national evidence linking women’s dietary diversity with underweight using age-appropriate anthropometric definitions remains limited.

Recent Cambodia-specific analyses of the same 2021–22 CDHS further illustrate the country’s changing nutrition and dietary profile among women of reproductive age. Um S et al., 2024 reported that 22.56% of non-pregnant women were overweight and 5.61% had obesity, with excess weight associated with factors including older age, higher household wealth, higher parity and alcohol consumption ^12^. A separate analysis found that 33.0% of women aged 15–49 years consumed unhealthy foods during the previous 24 hours; sweet foods, salty foods and sweet beverages were also commonly consumed, and unhealthy food consumption varied across sociodemographic groups ^13^. These studies provide important evidence on excess weight and unhealthy food consumption among Cambodian women, but they do not directly address whether achieving minimum dietary diversity is associated with underweight/thinness. This leaves a distinct evidence gap because MDD-W reflects food-group diversity and micronutrient adequacy rather than total energy intake, while underweight/thinness represents a different dimension of malnutrition. Examining these two measures together can therefore complement previous Cambodian evidence on unhealthy dietary practices and overweight/obesity and provide a more complete picture of the double burden of malnutrition among women.

Socioeconomic position is an important potential confounder in the relationship between women’s diet and nutritional status. Earlier Cambodian evidence found that household economic inequality was associated with women’s undernutrition ^14^. Cambodian research has also documented variation in women’s dietary diversity and nutritional status across socioeconomic and geographic groups ^15^. Studies in other low- and middle-income settings have similarly reported associations between household socioeconomic conditions, food security and women’s dietary diversity ^16^. Accordingly, household wealth quintile was treated as an a priori socioeconomic confounder in the present study and retained in the multivariable model regardless of its crude statistical significance.

Household wealth was retained a priori because socioeconomic position may influence both access to diverse foods and nutritional status through material living conditions, food affordability, disease exposure and access to services. This approach reduced reliance on statistical screening alone for an established socioeconomic confounder.

The policy relevance of women’s nutrition has also strengthened in Cambodia. The Third National Strategy for Food Security and Nutrition 2024–2028 recognises malnutrition as encompassing undernutrition, micronutrient deficiencies, overweight and obesity, and calls for healthier, nutrient-adequate diets and food-system actions that address all forms of malnutrition ^17^. This agenda is complemented by the Roadmap towards Universal Health Coverage in Cambodia 2024–2035, which links progress towards UHC to expanded social health protection, greater coverage of essential health services and improved financial risk protection ^18^. The Ministry of Health’s Health Strategic Plan 2025–2034 (HSP4), framed around effective and equitable health services for all through a resilient, responsive and accountable health system, aligns health-sector priorities with Cambodia’s 2030/2050 development vision and the Cambodia Sustainable Development Goals ^19^. Together, these national frameworks strengthen the rationale for population-level evidence that can identify nutritional inequalities among women and support integrated nutrition, prevention and health-service planning.

Therefore, this study aimed to estimate the prevalence and distribution of MDD-W and underweight/thinness and examine sociodemographic and dietary factors associated with underweight/thinness among Cambodian women aged 15–49 years.

## METHODS

### Study design and data source

This was a secondary cross-sectional analysis of the 2021–22 Cambodia Demographic and Health Survey (CDHS) ^11^. The CDHS used a stratified, multistage cluster sampling design to produce population-representative estimates ^11^. The present analysis used the women’s Individual Recode file and incorporated the survey sampling structure in estimation.

### Study population

The target population was women aged 15–49 years. Women who were currently pregnant were excluded because pregnancy changes body weight and makes conventional BMI-based nutritional classification inappropriate. Women who had given birth in the preceding 2 months were also excluded from the anthropometric analysis. Participants were required to have valid age-appropriate anthropometric information and complete information for constructing MDD-W.

The source IR file contained 19,496 interviewed women aged 15–49 years. **Figure 1** shows the sample-selection process. After excluding women who were currently pregnant, women who had given birth in the preceding 2 months, women without valid age-appropriate anthropometry, and women with incomplete MDD-W data, the final analytic sample comprised 9,181 women. The sum of survey weights in this analytic sample was 9,262.5; this weighted N is an analytic-sample measure and not a projected national population total.

**Figure 1.**
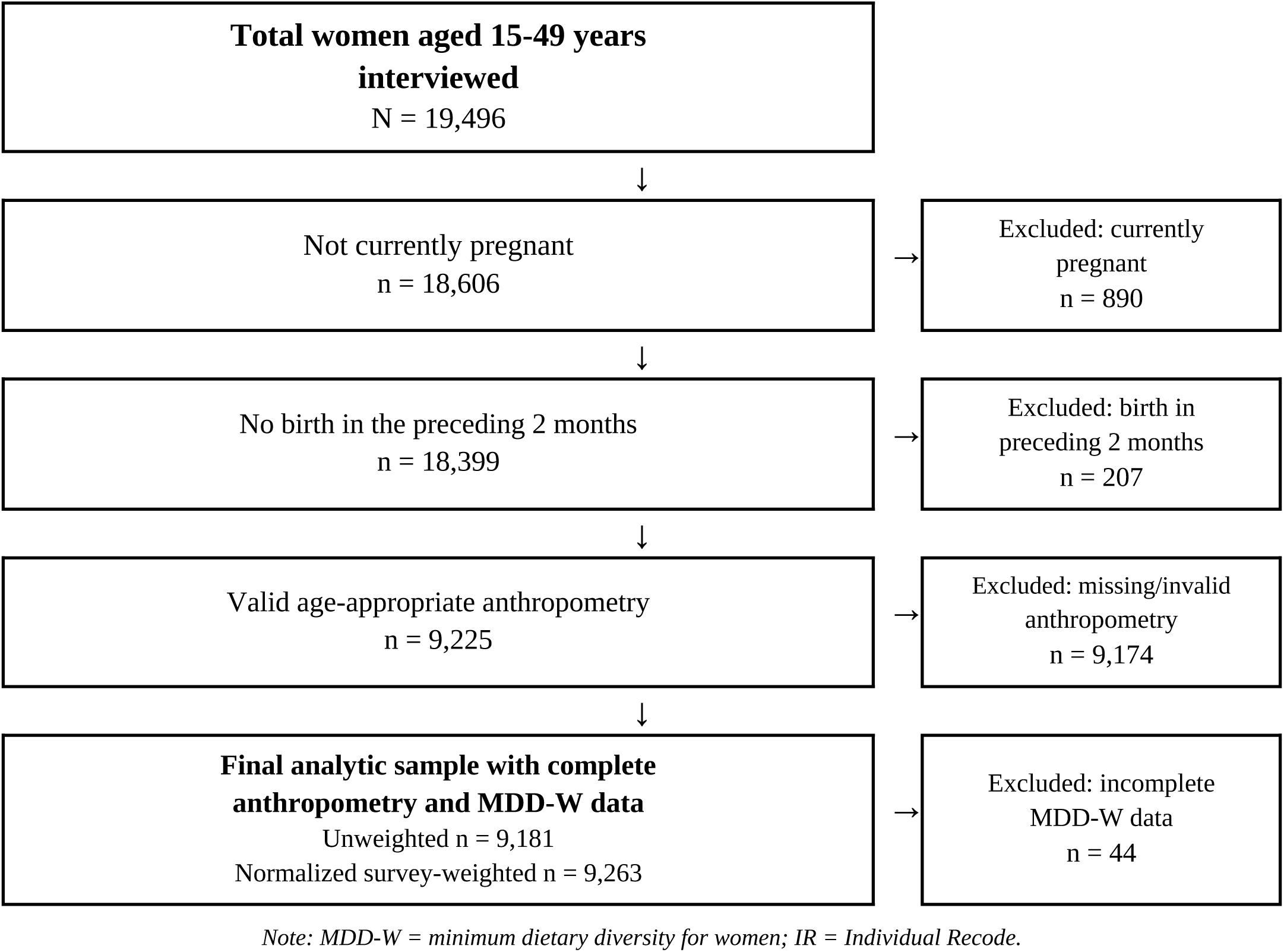
Study population flow diagram. MDD-W, minimum dietary diversity for women; IR, Individual Recode.

### Outcome: underweight/thinness and nutritional status

The primary outcome was underweight/thinness. Because the study included adolescents and adults, age-appropriate anthropometric definitions were used. For women aged 15–19 years, thinness was classified using the WHO BMI-for-age reference ^20^; for women aged 20–49 years, underweight was defined as BMI <18.5 kg/m² ^21^. For descriptive presentation of the full nutritional-status distribution, participants were classified as underweight/thinness, normal weight, overweight or obesity using age-appropriate adolescent and adult thresholds ^20 21^.

### Exposure: minimum dietary diversity for women

The primary exposure was MDD-W, constructed according to the ten-food-group framework used for women of reproductive age ^5–7^. Ten food groups were constructed from previous-day food-consumption items: (1) grains, white roots and tubers, and plantains; (2) pulses; (3) nuts and seeds; (4) milk and milk products; (5) meat, poultry and fish; (6) eggs; (7) dark green leafy vegetables; (8) other vitamin A-rich fruits and vegetables; (9) other vegetables; and (10) other fruits. One point was assigned for consumption of each group, and women consuming at least five of the ten groups were classified as achieving MDD-W ^5–7^. The ≥5-food-group cutoff was developed from multicountry dietary data as a practical population-level indicator related to micronutrient adequacy ^6 8^.

The ten MDD-W groups were reconstructed from the CDHS previous-day food-consumption items. Consistent with the CDHS report, the dairy group represents milk and milk products, and special or non-substantive response codes were treated as missing when constructing food-group indicators.

### Covariates

The primary exposure, MDD-W, was retained in the multivariable model irrespective of its crude association with underweight/thinness. Household wealth quintile was retained a priori as a socioeconomic confounder regardless of its crude P value because previous Cambodian and international evidence indicates that socioeconomic position is related to women’s dietary diversity, food security and undernutrition ^14–16^. Other candidate covariates included age group, residence, geographical region, education, marital status, current work status, parity, household size, travel time to the nearest health facility, household motorcycle/scooter ownership, household car/truck ownership, drinking-water source, toilet facility type, current smoking and alcohol use. These additional variables were considered for the adjusted model when their overall crude survey-weighted Wald P value was <0.20. For categorical predictors, selection was based on the overall variable-level test and all categories were retained when the variable met the inclusion criterion. Variables were not removed from the final model solely because their adjusted P value exceeded 0.05.

Covariates were grouped as demographic/life-stage, socioeconomic, household, geographic/access, WASH, and behavioural factors. Wealth quintile was retained regardless of its crude P value because it may influence both dietary diversity and nutritional status. For other categorical variables, entry into the multivariable model was based on the overall variable-level Wald test rather than category-specific P values, and all levels were retained when a variable met the criterion.

### Statistical analysis

All analyses incorporated the DHS sampling weight (v005/1,000,000), primary sampling unit (v021) and sampling stratum (v023), with the analytic sample treated as a survey domain within the full survey design. Unweighted sample sizes and normalized survey-weighted counts were reported where appropriate. Survey-weighted descriptive statistics were used to estimate participant characteristics, MDD-W, food-group consumption and nutritional status. Direct survey-weighted provincial estimates of MDD-W and underweight/thinness were visualised using choropleth maps without spatial interpolation or smoothing. Rao–Scott chi-square tests assessed differences in underweight/thinness across participant characteristics. Survey-weighted simple and multivariable logistic regression models estimated crude odds ratios (ORs) and adjusted odds ratios (aORs) with 95% confidence intervals. MDD-W and household wealth quintile were retained a priori in the adjusted model; other candidate covariates with an overall crude Wald P<0.20 were included in the multivariable model. Statistical significance in the final adjusted model was assessed at P<0.05. Multicollinearity in the final multivariable model was assessed using generalized variance inflation factors (GVIFs); for multi-degree-of-freedom categorical predictors, GVIF^(1/(2×df)) was used to facilitate comparison across terms (**Supplementary Table S6**). As a sensitivity analysis, the final multivariable survey-weighted logistic regression model was repeated among women aged 20–49 years only, using adult underweight defined uniformly as BMI <18.5 kg/m². The same survey design and final covariate set were retained to assess whether the primary MDD-W association was robust to exclusion of adolescents (**Supplementary Table S5**).

MDD-W was treated as the primary dietary exposure. Its ten component food groups were examined in separate secondary models rather than entered simultaneously with the MDD-W composite. Food-group prevalence and secondary food-group regression results are presented in Supplementary Tables S1 and S3, respectively. Provincial survey-weighted estimates of MDD-W and underweight/thinness were linked to Cambodia administrative boundaries and displayed as choropleth maps (**Figure 2**), with corresponding provincial estimates and 95% confidence intervals in **Supplementary Table S2**. These are direct provincial estimates; no spatial interpolation or smoothing was applied.

**Figure 2.**
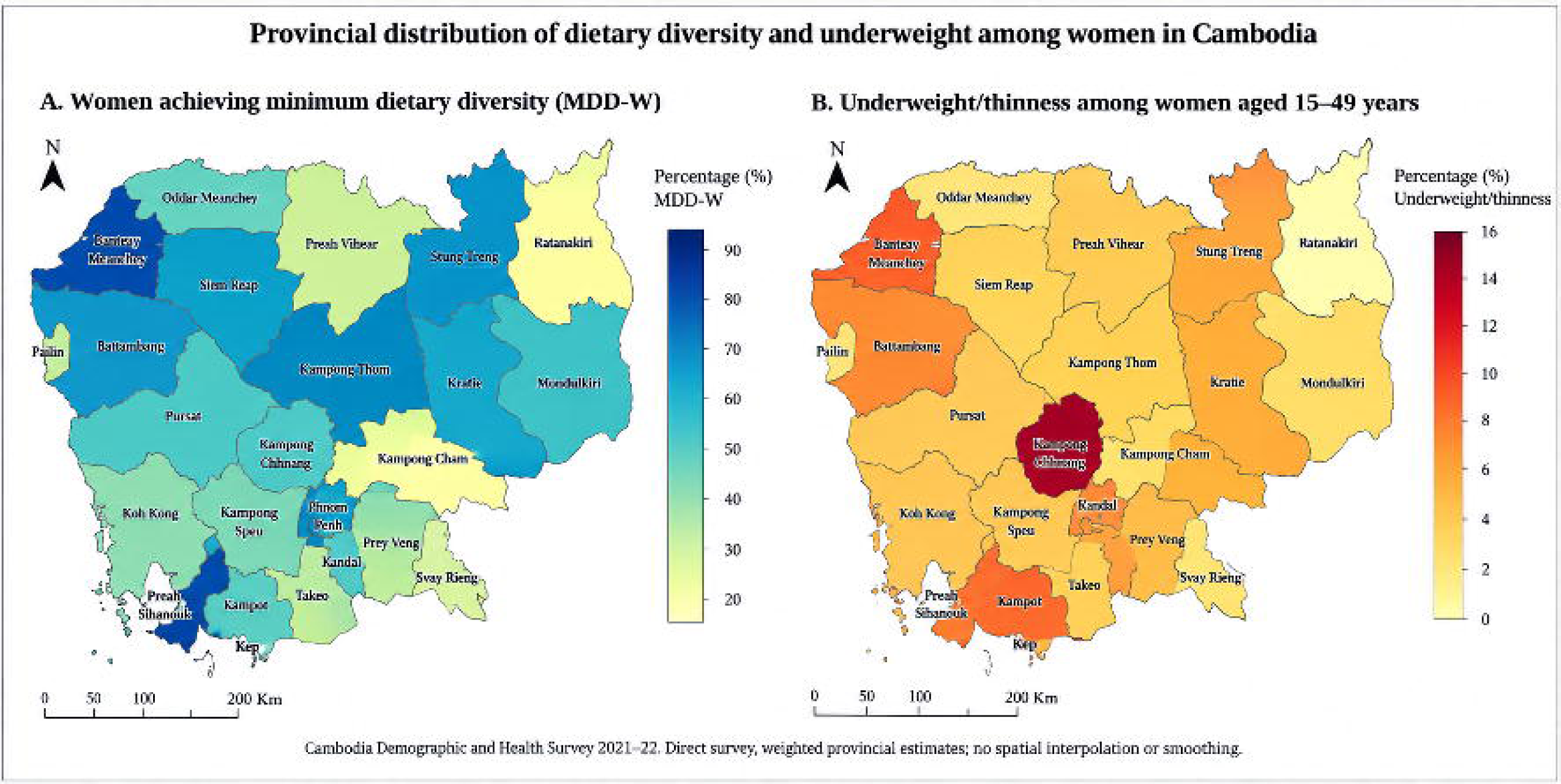
Provincial distribution of (**A**) minimum dietary diversity (MDD-W) and (**B**) underweight/thinness among women aged 15–49 years in Cambodia, 2021–22. Values are direct survey-weighted provincial estimates from the Cambodia Demographic and Health Survey 2021–22; no spatial interpolation or smoothing was applied.

Anthropometric measurements of women aged 15–49 years were collected in the half-sample of households not selected for the male survey. The analysis therefore used the CDHS survey design variables and the women’s individual sampling weight consistently across descriptive and regression analyses.

### Ethical considerations

The 2021–22 CDHS protocol was reviewed and approved by the Cambodia National Ethics Committee for Health Research and the ICF Institutional Review Board. Written informed consent was obtained in the original survey, with parental or guardian consent for respondents younger than 18 years. The present study used de-identified secondary data obtained through the DHS Program and required no new participant recruitment or contact.

### Patient and public involvement

This study was a secondary analysis of an existing national survey. No participants were newly recruited and there was no direct patient or public involvement in the design of this secondary analysis.

## RESULTS

### Study population

The final analytic sample comprised 9,181 women (weighted n= 9,262.5). **Figure 1** presents the sample-selection process, and **Table 1** summarizes survey-weighted participant characteristics.

**Table 1.** Survey-weighted characteristics of women aged 15–49 years, Cambodia DHS 2021–22.

| Characteristic |  | Weighted N | Weighted % (95% CI) |
| --- | --- | --- | --- |
| **Minimum dietary diversity (MDD-W)** |  |  |  |
|  | <5 food groups | 3,960.4 | 42.8 (41.0–44.5) |
|  | ≥5 food groups | 5,302.1 | 57.2 (55.5–59.0) |
| **Age group (years)** |  |  |  |
|  | 15–19 | 1,469.8 | 15.9 (14.9–16.8) |
|  | 20–24 | 1,131.2 | 12.2 (11.3–13.1) |
|  | 25–29 | 1,358.5 | 14.7 (13.3–16.0) |
|  | 30–34 | 1512 | 16.3 (15.1–17.5) |
|  | 35–39 | 1,639.9 | 17.7 (16.6–18.8) |
|  | 40–44 | 1293 | 14.0 (13.0–14.9) |
|  | 45–49 | 858.1 | 9.3 (8.5–10.1) |
| **Residence** |  |  |  |
|  | Urban | 3,950.4 | 42.6 (40.4–44.9) |
|  | Rural | 5,312.1 | 57.4 (55.1–59.6) |
| **Geographical region** |  |  |  |
|  | Phnom Penh | 1,531.1 | 16.5 (14.1–19.0) |
|  | Plains | 3,086.3 | 33.3 (31.6–35.0) |
|  | Tonle Sap | 2,674.3 | 28.9 (27.0–30.8) |
|  | Coastal/Sea | 581.2 | 6.3 (5.8–6.7) |
|  | Mountains/Plateau | 1,389.6 | 15.0 (14.1–15.9) |
| **Education** |  |  |  |
|  | No education | 1091 | 11.8 (10.8–12.8) |
|  | Primary | 3,603.9 | 38.9 (37.1–40.7) |
|  | Secondary | 3,909.4 | 42.2 (40.5–43.9) |
|  | Higher | 658.2 | 7.1 (6.0–8.2) |
| **Household wealth quintile** |  |  |  |
|  | Poorest | 1,583.8 | 17.1 (15.6–18.6) |
|  | Poorer | 1750 | 18.9 (17.3–20.4) |
|  | Middle | 1,791.2 | 19.3 (17.9–20.7) |
|  | Richer | 1,981.3 | 21.4 (19.8–23.0) |
|  | Richest | 2,156.1 | 23.3 (21.0–25.6) |
| **Marital status** |  |  |  |
|  | Never married | 2,414.7 | 26.1 (24.7–27.4) |
|  | Married/living together | 6,256.1 | 67.5 (66.1–68.9) |
|  | Formerly married | 591.7 | 6.4 (5.7–7.0) |
| **Currently working** |  |  |  |
|  | No | 3,089.4 | 33.4 (31.8–34.9) |
|  | Yes | 6,173.1 | 66.6 (65.1–68.2) |
| **Parity** |  |  |  |
|  | 0 | 2,762.2 | 29.8 (28.4–31.2) |
|  | 1–2 | 3,929.6 | 42.4 (40.8–44.1) |
|  | 3–4 | 2154 | 23.3 (22.1–24.5) |
|  | ≥5 | 416.7 | 4.5 (4.0–5.0) |
| **Household size** |  |  |  |
|  | ≤4 | 4,421.8 | 47.7 (45.8–49.7) |
|  | 5–6 | 3407 | 36.8 (35.1–38.5) |
|  | ≥7 | 1,433.7 | 15.5 (14.0–17.0) |
| **Travel time to nearest health facility** |  |  |  |
|  | <20 min | 6,092.8 | 65.8 (63.3–68.3) |
|  | 20–39 min | 2,677.3 | 28.9 (26.7–31.1) |
|  | ≥40 min | 492.4 | 5.3 (4.3–6.3) |
| **Household motorcycle/scooter ownership** |  |  |  |
|  | No | 920.7 | 10.0 (9.0–10.9) |
|  | Yes | 8,294.9 | 90.0 (89.1–91.0) |
| **Household car/truck ownership** |  |  |  |
|  | No | 7,491.2 | 81.3 (79.4–83.2) |
|  | Yes | 1,724.4 | 18.7 (16.8–20.6) |
| **Drinking-water source type** |  |  |  |
|  | Unimproved/other source type | 861.4 | 9.3 (8.1–10.6) |
|  | Improved source type | 8,354.2 | 90.7 (89.4–91.9) |
| **Toilet facility type** |  |  |  |
|  | Unimproved/no facility | 907.6 | 9.8 (8.6–11.1) |
|  | Improved facility type | 8308 | 90.2 (88.9–91.4) |
| **Current cigarette smoking** |  |  |  |
|  | No | 9,131.6 | 98.6 (98.3–98.9) |
|  | Yes | 130.8 | 1.4 (1.1–1.7) |
| **Alcohol consumption in past month** |  |  |  |
|  | No | 7,718.6 | 83.3 (82.1–84.5) |
|  | Yes | 1,543.9 | 16.7 (15.5–17.9) |

*All N values and percentages are survey-weighted. DHS weights are normalized; weighted N represents the weighted analytic sample rather than a projected national population total. MDD-W, minimum dietary diversity for women.*

### Nutritional status and dietary diversity

Overall, the survey-weighted prevalence of underweight/thinness was 6.7% (weighted cases approximately 616.6) (**Table 1**). The full nutritional-status distribution was 6.7% underweight/thinness, 64.9% normal weight, 22.8% overweight and 5.7% obesity (**Table 1**). Thus, excess weight was substantially more common than underweight/thinness in the overall analytic population, although nutritional status varied across age groups.

After weighting, approximately 5,302.1 women achieved MDD-W and 3,960.4 did not; 57.2% therefore consumed at least five of the ten MDD-W food groups, while 42.8% consumed fewer than five (**Table 1**). Starchy staples (99.0%) and flesh foods (96.7%) were the most commonly consumed food groups. Dark green leafy vegetables were consumed by 65.4%, other vegetables by 54.0%, other fruits by 51.0%, eggs by 42.3%, vitamin A-rich fruits and vegetables by 39.0%, dairy by 30.0%, pulses by 11.4%, and nuts and seeds by 6.9% (**Supplementary Table S1**). All percentages are survey-weighted.

### Provincial distribution

Provincial estimates showed geographic heterogeneity in both MDD-W and underweight/thinness (**Figure 2; Supplementary Table S2**). **Figure 2** presents direct survey-weighted provincial estimates and does not use spatial smoothing or interpolation. **Supplementary Table S2** provides the corresponding provincial estimates with 95% confidence intervals, which should be considered because precision varies with provincial sample size.

### Prevalence of underweight/thinness across participant characteristics

Underweight/thinness varied strongly by age in the current descriptive estimates (**Table 2**). Prevalence was 4.3% among women aged 15–19 years, increased to 17.7% among those aged 20–24 years, and was 9.4% among women aged 25–29 years. Prevalence was lower among older age groups: 4.2% at 30–34 years, 4.8% at 35–39 years, 3.6% at 40–44 years, and 4.3% at 45–49 years. Rural and urban prevalence estimates were similar (6.8% and 6.4%, respectively) (**Table 2**).

**Table 2.** Survey-weighted prevalence of underweight/thinness by participant characteristics.

| Characteristic |  | Not underweight/thinness weighted N (%) | Underweight/thinness weighted N (%) | P value |
| --- | --- | --- | --- | --- |
| Minimum dietary diversity (MDD-W) |  |  |  |  |
|  | <5 food groups | 3686.8 (93.1%) | 273.6 (6.9%) | 0.545 |
|  | ≥5 food groups | 4959.0 (93.5%) | 343.0 (6.5%) |  |
| Age group (years) |  |  |  |  |
|  | 15–19 | 1406.3 (95.7%) | 63.5 (4.3%) | <0.001 |
|  | 20–24 | 931.4 (82.3%) | 199.9 (17.7%) |  |
|  | 25–29 | 1230.2 (90.6%) | 128.3 (9.4%) |  |
|  | 30–34 | 1448.5 (95.8%) | 63.6 (4.2%) |  |
|  | 35–39 | 1561.8 (95.2%) | 78.1 (4.8%) |  |
|  | 40–44 | 1246.9 (96.4%) | 46.1 (3.6%) |  |
|  | 45–49 | 820.9 (95.7%) | 37.2 (4.3%) |  |
| Residence |  |  |  |  |
|  | Urban | 3696.4 (93.6%) | 254.0 (6.4%) | 0.602 |
|  | Rural | 4949.5 (93.2%) | 362.6 (6.8%) |  |
| Geographical region |  |  |  |  |
|  | Phnom Penh | 1430.2 (93.4%) | 100.9 (6.6%) | 0.138 |
|  | Plains | 2902.8 (94.1%) | 183.5 (5.9%) |  |
|  | Tonle Sap | 2468.6 (92.3%) | 205.7 (7.7%) |  |
|  | Coastal/Sea | 531.3 (91.4%) | 49.9 (8.6%) |  |
|  | Mountains/Plateau | 1313.0 (94.5%) | 76.6 (5.5%) |  |
| Education |  |  |  |  |
|  | No education | 1040.6 (95.4%) | 50.5 (4.6%) | <0.001 |
|  | Primary | 3386.3 (94.0%) | 217.6 (6.0%) |  |
|  | Secondary | 3673.7 (94.0%) | 235.6 (6.0%) |  |
|  | Higher | 545.3 (82.9%) | 112.9 (17.1%) |  |
| Household wealth quintile |  |  |  |  |
|  | Poorest | 1466.0 (92.6%) | 117.8 (7.4%) | 0.619 |
|  | Poorer | 1623.7 (92.8%) | 126.4 (7.2%) |  |
|  | Middle | 1680.0 (93.8%) | 111.2 (6.2%) |  |
|  | Richer | 1863.0 (94.0%) | 118.4 (6.0%) |  |
|  | Richest | 2013.2 (93.4%) | 142.9 (6.6%) |  |
| Marital status |  |  |  |  |
|  | Never married | 2152.2 (89.1%) | 262.5 (10.9%) | <0.001 |
|  | Married/living together | 5934.4 (94.9%) | 321.7 (5.1%) |  |
|  | Formerly married | 559.3 (94.5%) | 32.4 (5.5%) |  |
| Currently working |  |  |  |  |
|  | No | 2863.9 (92.7%) | 225.5 (7.3%) | 0.154 |
|  | Yes | 5782.0 (93.7%) | 391.1 (6.3%) |  |
| Parity |  |  |  |  |
|  | 0 | 2463.3 (89.2%) | 298.9 (10.8%) | <0.001 |
|  | 1–2 | 3710.2 (94.4%) | 219.4 (5.6%) |  |
|  | 3–4 | 2078.9 (96.5%) | 75.1 (3.5%) |  |
|  | ≥5 | 393.5 (94.4%) | 23.2 (5.6%) |  |
| Household size |  |  |  |  |
|  | ≤4 | 4147.3 (93.8%) | 274.5 (6.2%) | 0.025 |
|  | 5–6 | 3190.3 (93.6%) | 216.7 (6.4%) |  |
|  | ≥7 | 1308.2 (91.2%) | 125.5 (8.8%) |  |
| Travel time to nearest health facility |  |  |  |  |
|  | <20 min | 5684.1 (93.3%) | 408.7 (6.7%) | 0.963 |
|  | 20–39 min | 2501.9 (93.4%) | 175.4 (6.6%) |  |
|  | ≥40 min | 459.8 (93.4%) | 32.5 (6.6%) |  |
| Household motorcycle/scooter ownership |  |  |  |  |
|  | No | 846.3 (91.9%) | 74.4 (8.1%) | 0.141 |
|  | Yes | 7754.4 (93.5%) | 540.5 (6.5%) |  |
| Household car/truck ownership |  |  |  |  |
|  | No | 6965.5 (93.0%) | 525.7 (7.0%) | 0.048 |
|  | Yes | 1635.1 (94.8%) | 89.3 (5.2%) |  |
| Drinking-water source type |  |  |  |  |
|  | Unimproved/other source type | 785.9 (91.2%) | 75.5 (8.8%) | 0.043 |
|  | Improved source type | 7814.7 (93.5%) | 539.4 (6.5%) |  |
| Toilet facility type |  |  |  |  |
|  | Unimproved/no facility | 847.7 (93.4%) | 59.9 (6.6%) | 0.934 |
|  | Improved facility type | 7753.0 (93.3%) | 555.0 (6.7%) |  |
| Current cigarette smoking |  |  |  |  |
|  | No | 8526.5 (93.4%) | 605.2 (6.6%) | 0.372 |
|  | Yes | 119.4 (91.2%) | 11.4 (8.8%) |  |
| Alcohol consumption in past month |  |  |  |  |
|  | No | 7183.6 (93.1%) | 535.0 (6.9%) | 0.108 |
|  | Yes | 1462.2 (94.7%) | 81.6 (5.3%) |  |

N and percentages are survey-weighted; percentages are row percentages. Overall P values are design-adjusted Rao–Scott tests.

**Table 2** also shows variation in underweight/thinness prevalence by education and other participant characteristics. These differences should be interpreted together with the age distribution and the multivariable estimates in **Table 3** because several characteristics are strongly related to age and life stage.

**Table 3.** Survey-weighted crude and adjusted associations with underweight/thinness.

| Characteristic |  | Crude OR (95% CI) | P value | Adjusted OR (95% CI) | P value |
| --- | --- | --- | --- | --- | --- |
| **Minimum dietary diversity (MDD-W)** |  |  |  |  |  |
|  | <5 food groups | 1.00 (reference) |  | 1.00 (reference) |  |
|  | ≥5 food groups | 0.93 (0.74–1.17) | 0.545 | 0.86 (0.67–1.09) | 0.219 |
| **Age group (years)** |  |  |  |  |  |
|  | 15–19 | 1.00 (reference) |  | 1.00 (reference) |  |
|  | 20–24 | 4.75 (3.41–6.62) | <0.001 | 6.40 (4.39–9.34) | <0.001 |
|  | 25–29 | 2.31 (1.56–3.42) | <0.001 | 4.35 (2.72–6.96) | <0.001 |
|  | 30–34 | 0.97 (0.63–1.49) | 0.896 | 2.24 (1.30–3.84) | 0.004 |
|  | 35–39 | 1.11 (0.72–1.71) | 0.645 | 2.86 (1.66–4.92) | <0.001 |
|  | 40–44 | 0.82 (0.51–1.30) | 0.398 | 2.17 (1.21–3.90) | 0.010 |
|  | 45–49 | 1.00 (0.61–1.66) | 0.985 | 2.59 (1.39–4.83) | 0.003 |
| **Residence** |  |  |  |  |  |
|  | Urban | 1.00 (reference) |  | — | — |
|  | Rural | 1.07 (0.84–1.36) | 0.602 | — | — |
| **Geographical region** |  |  |  |  |  |
|  | Phnom Penh | 1.00 (reference) |  | 1.00 (reference) |  |
|  | Plains | 0.90 (0.58–1.38) | 0.618 | 0.91 (0.51–1.62) | 0.743 |
|  | Tonle Sap | 1.18 (0.77–1.82) | 0.449 | 1.07 (0.61–1.88) | 0.813 |
|  | Coastal/Sea | 1.33 (0.79–2.24) | 0.277 | 1.34 (0.72–2.52) | 0.360 |
|  | Mountains/Plateau | 0.83 (0.53–1.30) | 0.407 | 0.77 (0.42–1.42) | 0.401 |
| **Education** |  |  |  |  |  |
|  | No education | 1.00 (reference) |  | 1.00 (reference) |  |
|  | Primary | 1.32 (0.93–1.89) | 0.121 | 1.26 (0.88–1.81) | 0.202 |
|  | Secondary | 1.32 (0.93–1.88) | 0.117 | 1.08 (0.73–1.60) | 0.711 |
|  | Higher | 4.27 (2.80–6.50) | <0.001 | 2.48 (1.45–4.23) | <0.001 |
| **Household wealth quintile** |  |  |  |  |  |
|  | Poorest | 1.00 (reference) |  | 1.00 (reference) |  |
|  | Poorer | 0.97 (0.72–1.30) | 0.831 | 0.93 (0.65–1.33) | 0.694 |
|  | Middle | 0.82 (0.61–1.12) | 0.212 | 0.76 (0.54–1.09) | 0.138 |
|  | Richer | 0.79 (0.57–1.11) | 0.171 | 0.82 (0.55–1.22) | 0.329 |
|  | Richest | 0.88 (0.62–1.25) | 0.487 | 0.77 (0.46–1.28) | 0.308 |
| **Marital status** |  |  |  |  |  |
|  | Never married | 1.00 (reference) |  | 1.00 (reference) |  |
|  | Married/living together | 0.44 (0.36–0.54) | <0.001 | 0.78 (0.47–1.29) | 0.331 |
|  | Formerly married | 0.48 (0.31–0.74) | <0.001 | 0.77 (0.42–1.42) | 0.398 |
| **Currently working** |  |  |  |  |  |
|  | No | 1.00 (reference) |  | 1.00 (reference) |  |
|  | Yes | 0.86 (0.70–1.06) | 0.155 | 0.82 (0.65–1.03) | 0.093 |
| **Parity** |  |  |  |  |  |
|  | 0 | 1.00 (reference) |  | 1.00 (reference) |  |
|  | 1–2 | 0.49 (0.40–0.60) | <0.001 | 0.55 (0.35–0.86) | 0.010 |
|  | 3–4 | 0.30 (0.21–0.42) | <0.001 | 0.39 (0.23–0.69) | 0.001 |
|  | ≥5 | 0.49 (0.29–0.80) | 0.005 | 0.57 (0.28–1.16) | 0.119 |
| **Household size** |  |  |  |  |  |
|  | ≤4 | 1.00 (reference) |  | 1.00 (reference) |  |
|  | 5–6 | 1.03 (0.82–1.29) | 0.823 | 1.20 (0.94–1.53) | 0.146 |
|  | ≥7 | 1.45 (1.09–1.93) | 0.011 | 1.48 (1.09–2.01) | 0.013 |
| **Travel time to nearest health facility** |  |  |  |  |  |
|  | <20 min | 1.00 (reference) |  | — | — |
|  | 20–39 min | 0.98 (0.76–1.25) | 0.841 | — | — |
|  | ≥40 min | 0.98 (0.69–1.41) | 0.931 | — | — |
| **Household motorcycle/scooter ownership** |  |  |  |  |  |
|  | No | 1.00 (reference) |  | 1.00 (reference) |  |
|  | Yes | 0.79 (0.58–1.08) | 0.142 | 0.76 (0.55–1.05) | 0.094 |
| **Household car/truck ownership** |  |  |  |  |  |
|  | No | 1.00 (reference) |  | 1.00 (reference) |  |
|  | Yes | 0.72 (0.52–1.00) | 0.049 | 0.64 (0.43–0.94) | 0.022 |
| **Drinking-water source type** |  |  |  |  |  |
|  | Unimproved/other source type | 1.00 (reference) |  | 1.00 (reference) |  |
|  | Improved source type | 0.72 (0.52–0.99) | 0.044 | 0.75 (0.52–1.09) | 0.133 |
| **Toilet facility type** |  |  |  |  |  |
|  | Unimproved/no facility | 1.00 (reference) |  | — | — |
|  | Improved facility type | 1.01 (0.75–1.38) | 0.934 | — | — |
| **Current cigarette smoking** |  |  |  |  |  |
|  | No | 1.00 (reference) |  | — | — |
|  | Yes | 1.35 (0.70–2.63) | 0.374 | — | — |
| **Alcohol consumption in past month** |  |  |  |  |  |
|  | No | 1.00 (reference) |  | 1.00 (reference) |  |
|  | Yes | 0.75 (0.53–1.07) | 0.110 | 0.76 (0.53–1.09) | 0.136 |

MDD-W and household wealth quintile were retained a priori in the adjusted model. Other candidate variables entered the model when the overall crude survey-weighted Wald $P < 0.20$ . OR, odds ratio; aOR, adjusted odds ratio; CI, confidence interval.

### Association between MDD-W and underweight/thinness

In crude survey-weighted logistic regression, achieving MDD-W was not associated with underweight/thinness (OR 0.93, 95% CI 0.74 to 1.17; P=0.545) (**Table 3**). Household wealth quintile was retained a priori in the adjusted model regardless of its crude P value. In the adjusted survey-weighted model, the association between MDD-W and underweight/thinness remained imprecise (aOR 0.86, 95% CI 0.67 to 1.09; P=0.219).

Age showed the strongest association with underweight/thinness in the adjusted estimates (**Table 3**). Compared with women aged 15–19 years, women aged 20–24 years had substantially higher odds of underweight/thinness (aOR 6.40, 95% CI 4.39 to 9.34), and elevated odds were also observed in several older age groups. Interpretation should account for the age-appropriate outcome definition: WHO BMI-for-age thinness was used at ages 15–19 years and adult BMI-defined underweight from age 20 years onward.

### Individual food groups

#### Adult-only sensitivity analysis

In the sensitivity analysis restricted to women aged 20–49 years, achieving MDD-W remained not clearly associated with adult underweight after adjustment (aOR 0.89, 95% CI 0.68 to 1.17; P=0.401) (**Supplementary Table S5**). This was consistent with the direction and interpretation of the primary analysis that included women aged 15–49 years.

In secondary models examining individual MDD-W food groups separately (**Supplementary Table S3**), most associations had confidence intervals that included the null. Consumption of other vegetables was associated with lower odds of underweight/thinness (aOR 0.78, 95% CI 0.61 to 1.00; P=0.047), while the estimate for vitamin A-rich fruits and vegetables was in the same direction but imprecise (aOR 0.81, 95% CI 0.64 to 1.01; P=0.067). These secondary analyses are exploratory because multiple food groups were examined.

## DISCUSSION

### Key findings

This nationally representative analysis highlights three related features of women’s nutrition in Cambodia. First, 57.2% of women achieved MDD-W. Second, 6.7% had underweight/thinness, while 22.8% were overweight and 5.7% had obesity, demonstrating the coexistence of undernutrition and excess weight. Third, neither the crude nor the adjusted estimate showed clear evidence of an association between achieving MDD-W and underweight/thinness. MDD-W was retained as the primary exposure and household wealth quintile as an a priori socioeconomic confounder.

The absence of a clear independent association between MDD-W and underweight/thinness should not be interpreted as evidence that dietary diversity is unimportant. MDD-W was developed as a population-level indicator of food-group diversity and micronutrient adequacy, rather than as a measure of total energy intake, portion size or energy balance ^5–8^. Validation studies support its usefulness for identifying diets with greater micronutrient adequacy, while also showing that the method of dietary assessment can influence classification ^8 9^. Anthropometric underweight, by contrast, reflects the cumulative balance of energy intake and expenditure together with health, biological and socioeconomic influences. These measures therefore capture different dimensions of nutritional status, providing a plausible explanation for the null association in the primary model.

### Interpretation in the Cambodian context

The nutritional-status distribution observed in this analysis is consistent with Cambodia’s continuing nutrition transition and the broader double burden of malnutrition described in low- and middle-income countries ^1 3 4^. Earlier work using Cambodian DHS data documented declining underweight alongside increasing overweight among women and persistent socioeconomic inequalities in nutritional status ^10^. The current findings extend that context by showing that, among women with available anthropometry and dietary data in the 2021–22 CDHS, undernutrition remains present while overweight and obesity together affect more than one-quarter of the analytic population (**Table 1**). This coexistence argues for nutrition surveillance and programmes that consider both insufficient and excessive nutritional status rather than treating them as separate population problems.

Age was more strongly associated with underweight/thinness than MDD-W in the current regression output (**Table 3**). The particularly high descriptive prevalence among women aged 20–24 years warrants further investigation, but the cross-sectional design cannot establish the reasons for this pattern. Interpretation should also recognise the age-specific outcome definition: thinness among women aged 15–19 years is based on BMI-for-age, whereas underweight from age 20 years is based on the adult BMI threshold. The adult-only sensitivity analysis among women aged 20–49 years produced a similar MDD-W estimate (aOR 0.89, 95% CI 0.68 to 1.17; P=0.401), supporting the robustness of the primary null association to exclusion of adolescents (**Supplementary Table S5**).

Socioeconomic interpretation is equally important. Previous Cambodian studies have documented inequalities in women’s nutritional status, and the present modeling strategy therefore retains household wealth quintile regardless of its crude association with the outcome ^10^. Wealth can influence the affordability and availability of diverse foods and may also capture broader differences in housing, sanitation, transport, access to services and other material conditions. Adjustment for wealth is thus intended primarily to reduce socioeconomic confounding of the MDD-W association. The wealth coefficients themselves should be interpreted cautiously because wealth is an asset-based relative measure within the survey population and because several related household variables are also considered in the analysis. Multicollinearity diagnostics did not indicate substantial collinearity in the final model; the largest adjusted GVIF [GVIF^(1/(2×df))] was 1.50 (**Supplementary Table S6**). Individual socioeconomic coefficients should nevertheless be interpreted in the context of related household characteristics.

### Dietary patterns

The dietary profile provides additional context for the MDD-W result. Consumption of starchy staples and flesh foods was very common, whereas pulses and nuts and seeds were much less frequently consumed (**Supplementary Table S1**). Greater food-group diversity has been associated with greater micronutrient adequacy in multicountry validation studies ^6 8^, but the MDD-W indicator does not quantify how much food was consumed ^5–7^. Thus, women can achieve the five-food-group threshold without necessarily meeting energy requirements, while women with lower dietary diversity may still consume sufficient energy. The individual food-group models in **Supplementary Table S3** should consequently be viewed as exploratory. The borderline association observed for other vegetables should not be interpreted in isolation because several food groups were tested and the confidence interval approached the null.

The high prevalence of starchy staples and flesh foods, contrasted with the low prevalence of pulses and nuts and seeds, also shows that identical MDD-W scores can arise from different food-group combinations. Component food-group models were therefore treated as secondary analyses and isolated borderline P values were not interpreted as definitive evidence. Future work using repeated or quantitative dietary assessment could examine whether specific dietary patterns, portion sizes or nutrient intakes are more closely related to low BMI than the dichotomous MDD-W indicator.

### Strengths and limitations

The study has several strengths. It uses data from a large national household survey, applies the survey weighting and clustered and stratified sampling structure, and uses age-appropriate anthropometric definitions for adolescents and adults. The analysis uses the established ten-food-group MDD-W framework and separates the primary composite MDD-W analysis from secondary food-group analyses. The modeling strategy also prespecifies MDD-W as the primary exposure and household wealth quintile as an a priori socioeconomic confounder, while using an overall variable-level Wald criterion for additional covariate selection. The adult-only sensitivity analysis further assessed robustness to the use of different age-appropriate anthropometric definitions for adolescents and adults. Several limitations are important. First, the cross-sectional design prevents determination of temporal sequence and therefore does not support causal inference. Second, MDD-W is based on previous-day food-group consumption and does not measure usual intake, total energy, protein intake or portion size ^5–7^; dietary recall and food-group classification error are also possible ^9^. Third, anthropometry was available only for the CDHS subsample in which women were measured, which reduced the analytic sample; all estimates therefore incorporated the DHS women’s individual sampling weight and the clustered and stratified survey design. Fourth, MDD-W was reconstructed from the available CDHS previous-day food-consumption recode items; the resulting national MDD-W estimate was consistent with the published CDHS report, but some food-group misclassification remains possible. Fifth, adolescent thinness and adult underweight use different age-appropriate definitions ^20 21^, limiting simple comparisons across the 19-to-20-year boundary; however, the adult-only sensitivity analysis yielded a similar primary MDD-W association. Sixth, residual confounding is possible because the survey does not capture every determinant of body weight or dietary intake. Finally, the individual food-group analyses involved multiple comparisons and should be considered secondary and hypothesis-generating.

### Implications for policy and research

The findings support a multidimensional approach to women’s nutrition in Cambodia. MDD-W can contribute useful information on food-group diversity, but it should be interpreted alongside anthropometry and other measures of dietary quantity, food security and socioeconomic conditions. The provincial variation shown in **Figure 2** may help identify areas for further assessment, although direct provincial estimates should always be interpreted with their confidence intervals. Future studies could combine dietary diversity with quantitative dietary assessment and examine effect modification by age and reproductive life stage. Longitudinal data would be particularly useful for clarifying temporal relationships between dietary patterns and subsequent nutritional status.

These findings are relevant to Cambodia’s current policy direction. The Third National Strategy for Food Security and Nutrition 2024–2028 addresses multiple forms of malnutrition, while the UHC Roadmap 2024–2035 and Health Strategic Plan 2025–2034 emphasise equitable access to essential services ^11 17–19^. Women’s nutrition surveillance can contribute by monitoring persistent undernutrition alongside overweight and obesity. The results support combining diet-quality indicators with anthropometry, socioeconomic information and service-access measures rather than assuming that increasing dietary diversity alone will reduce underweight.

## CONCLUSION

Among Cambodian women aged 15–49 years in the 2021–22 CDHS analytic sample, 57.2% achieved minimum dietary diversity and 6.7% had underweight/thinness, while 22.8% were overweight and 5.7% had obesity. Achieving MDD-W was not clearly associated with underweight/thinness. Dietary diversity and anthropometric status therefore capture related but distinct dimensions of women’s nutrition. Nutrition surveillance and future research should consider undernutrition and excess weight together and complement food-group diversity with measures of dietary quantity, household resources and life stage.

## Data Availability

The 2021-22 Cambodia Demographic and Health Survey data are available to registered users through the DHS Program subject to data-access approval and the DHS terms of use.

https://dhsprogram.com/data/available-datasets.cfm

## ACKNOWLEDGEMENTS

The authors acknowledge the Ministry of Health, the National Institute of Statistics, ICF, the survey field teams and all women who participated in the 2021–22 Cambodia Demographic and Health Survey.

## CONTRIBUTORS

**Chantrea Sieng, Samnang Um and Sovandara Heng** contributed equally to this paper. All authors contributed to the study conception and design, development of the analysis plan, interpretation of the findings, critical revision of the manuscript and approval of the final version. Samnang Um conducted the statistical analysis and prepared the initial manuscript draft. All authors accept responsibility for the integrity and accuracy of the work. Chantrea Sieng is the guarantor and accepts full responsibility for the overall content of the work.

## AI USE DECLARATION

OpenAI ChatGPT was used to assist with language editing, manuscript organization, and development/checking of analysis code. The authors remain responsible for verification of all AI-assisted outputs and for the final content of the manuscript.

## DATA AVAILABILITY STATEMENT

The 2021–22 Cambodia Demographic and Health Survey data are available to registered users through the DHS Program subject to data-access approval and the DHS terms of use.

## SUPPLEMENTARY FILES

**Supplementary Table S1**. Survey-weighted prevalence of the ten MDD-W food groups

**Supplementary Table S2**. Survey-weighted provincial estimates of MDD-W and underweight/thinness

**Supplementary Table S3**. Secondary individual food-group regression models

**Supplementary Table S4**. Candidate-variable screening and final-model rule

**Supplementary Table S5**. Adult-only sensitivity analysis among women aged 20–49 years

**Supplementary Table S6**. Multicollinearity diagnostics for the final multivariable model

