## Supplement for "Dietary Diversity and Underweight Among Women Aged 15–49 Years in Cambodia: Analysis of the 2021–22 Cambodia Demographic and Health Survey"

### **Supplementary Table S1.** Survey-weighted prevalence of the ten MDD-W food groups

| **Food group** | **Weighted women consuming** | **Weighted % (95% CI)** |
| --- | --- | --- |
| Grains, white roots/tubers and plantains | 9168 | 99.0 (98.7–99.2) |
| Pulses | 1,055.8 | 11.4 (10.3–12.5) |
| Nuts and seeds | 635.3 | 6.9 (6.0–7.7) |
| Dairy | 2,778.9 | 30.0 (28.3–31.7) |
| Meat, poultry and fish | 8,960.1 | 96.7 (96.2–97.3) |
| Eggs | 3,916.7 | 42.3 (40.8–43.7) |
| Dark green leafy vegetables | 6,054.9 | 65.4 (63.8–66.9) |
| Other vitamin A-rich fruits and vegetables | 3,609.7 | 39.0 (37.4–40.6) |
| Other vegetables | 5,005.5 | 54.0 (52.4–55.7) |
| Other fruits | 4,723.4 | 51.0 (49.5–52.5) |

All N values and percentages are survey-weighted.

### **Supplementary Table S2.** Survey-weighted provincial estimates of MDD-W and underweight/thinness

| **Province** | **Weighted women (N)** | **MDD-W % (95% CI)** | **Underweight/thinness % (95% CI)** |
| --- | --- | --- | --- |
| Banteay Meanchey | 360.1 | 67.6 (60.2–75.0) | 8.8 (5.8–11.8) |
| Battambang | 643.6 | 61.3 (56.2–66.3) | 7.6 (4.6–10.5) |
| Kampong Cham | 554.8 | 47.9 (42.1–53.6) | 4.1 (2.4–5.8) |
| Kampong Chhnang | 327.5 | 59.0 (53.0–64.9) | 16.9 (12.9–20.9) |
| Kampong Speu | 592.4 | 53.3 (47.2–59.3) | 6.2 (3.8–8.6) |
| Kampong Thom | 388.7 | 61.4 (54.5–68.3) | 5.4 (3.2–7.6) |
| Kampot | 373.3 | 49.2 (41.6–56.7) | 9.6 (5.6–13.7) |
| Kandal | 666.7 | 59.2 (53.0–65.4) | 7.1 (4.3–9.9) |
| Kep | 26.8 | 66.1 (60.6–71.6) | 6.6 (4.4–8.8) |
| Koh Kong | 66.5 | 50.8 (42.8–58.9) | 6.4 (3.0–9.8) |
| Kratie | 209.8 | 60.6 (54.7–66.5) | 7.6 (5.0–10.3) |
| Mondul Kiri | 54.5 | 56.9 (49.1–64.8) | 4.1 (2.6–5.5) |
| Otdar Meanchey | 112.7 | 50.5 (41.8–59.1) | 4.0 (1.9–6.0) |
| Pailin | 44.1 | 70.4 (65.0–75.7) | 2.7 (0.5–5.0) |
| Phnom Penh | 1,531.1 | 80.3 (76.2–84.4) | 6.6 (4.2–9.0) |
| Preah Sihanouk | 114.6 | 79.4 (74.6–84.1) | 6.9 (4.6–9.2) |
| Preah Vihear | 151.4 | 31.5 (24.5–38.6) | 5.6 (3.0–8.2) |
| Prey Veng | 567.4 | 49.3 (43.4–55.3) | 6.6 (4.0–9.2) |
| Pursat | 218.2 | 57.0 (50.7–63.4) | 5.3 (2.9–7.8) |
| Ratanakiri | 133.1 | 30.5 (24.3–36.7) | 0.7 (0.0–1.5) |
| Siemreap | 736.2 | 57.1 (48.4–65.7) | 5.1 (2.3–7.9) |
| Stung Treng | 91.4 | 61.2 (55.5–66.9) | 7.2 (4.4–10.0) |
| Svay Rieng | 349.9 | 34.5 (29.8–39.2) | 4.8 (2.4–7.2) |
| Takeo | 541.3 | 46.1 (39.1–53.1) | 5.8 (3.6–8.0) |
| Tboung Khmum | 406.2 | 28.7 (22.7–34.7) | 6.9 (4.0–9.7) |

Direct survey-weighted provincial estimates; no spatial interpolation or smoothing was applied. Confidence intervals should be considered when interpreting province-level estimates.

### **Supplementary Table S3**. Secondary individual food-group regression models

| **Food group** | **Weighted analytic N** | **aOR (95% CI)** | **P value** |
| --- | --- | --- | --- |
| Starchy staples | 9,262.5 | 0.54 (0.23-1.29) | 0.166 |
| Pulses | 9,262.5 | 1.11 (0.76-1.60) | 0.597 |
| Nuts and seeds | 9,262.5 | 0.92 (0.57-1.48) | 0.718 |
| Dairy | 9,262.5 | 0.92 (0.72-1.19) | 0.531 |
| Flesh foods | 9,262.5 | 1.27 (0.77-2.08) | 0.350 |
| Eggs | 9,262.5 | 1.12 (0.90-1.39) | 0.306 |
| Dark green leafy vegetables | 9,262.5 | 0.96 (0.76-1.22) | 0.765 |
| Vitamin A-rich fruits/vegetables | 9,262.5 | 0.81 (0.64-1.01) | 0.067 |
| Other vegetables | 9,262.5 | 0.78 (0.61-1.00) | 0.047 |
| Other fruits | 9,262.5 | 0.86 (0.69-1.06) | 0.152 |

Each food group was examined in a separate secondary model rather than simultaneously with MDD-W. Estimates are interpreted as exploratory because multiple food groups were examined.

### **Supplementary Table S4.** Candidate-variable screening and final-model rule

| **Candidate variable** | **Crude P value** | **Final model rule/status** |
| --- | --- | --- |
| MDD-W | 0.545 | Forced: primary exposure |
| Age group | <0.001 | Included |
| Residence | 0.602 | Not selected |
| Geographical region | 0.048 | Included |
| Education | <0.001 | Included |
| Household wealth quintile | 0.617 | Forced a priori socioeconomic confounder |
| Marital status | <0.001 | Included |
| Currently working | 0.155 | Included |
| Parity | <0.001 | Included |
| Household size | 0.032 | Included |
| Travel time to nearest health facility | 0.979 | Not selected |
| Household motorcycle/scooter ownership | 0.142 | Included |
| Household car/truck ownership | 0.049 | Included |
| Drinking-water source | 0.044 | Included |
| Toilet facility type | 0.934 | Not selected |
| Currently smokes | 0.374 | Not selected |
| Alcohol use in past month | 0.110 | Included |

*MDD-W and household wealth quintile are retained a priori regardless of crude P value; other candidate variables are considered when the overall crude survey-weighted Wald P<0.20.*

**Supplementary Table S5.** Adult-only sensitivity analysis among women aged 20–49 years

| **Characteristic** | | **Adjusted OR (95% CI)** | **P value** |
| --- | --- | --- | --- |
| **Minimum dietary diversity (MDD-W)** | |  |  |
|  | <5 food groups | 1.00 (reference) |  |
|  | ≥5 food groups | 0.89 (0.68–1.17) | 0.401 |
| **Household wealth quintile** | |  |  |
|  | Poorest | 1.00 (reference) |  |
|  | Poorer | 0.90 (0.62–1.29) | 0.560 |
|  | Middle | 0.68 (0.47–1.00) | 0.047 |
|  | Richer | 0.66 (0.43–1.02) | 0.060 |
|  | Richest | 0.61 (0.36–1.06) | 0.077 |
| **Age group (years)** | |  |  |
|  | 20–24 | 1.00 (reference) |  |
|  | 25–29 | 0.69 (0.49–0.98) | 0.039 |
|  | 30–34 | 0.36 (0.24–0.54) | <0.001 |
|  | 35–39 | 0.46 (0.29–0.71) | <0.001 |
|  | 40–44 | 0.35 (0.22–0.55) | <0.001 |
|  | 45–49 | 0.42 (0.25–0.69) | <0.001 |
| **Geographical region** | |  |  |
|  | Phnom Penh | 1.00 (reference) |  |
|  | Plains | 0.99 (0.55–1.79) | 0.985 |
|  | Tonle Sap | 1.06 (0.60–1.89) | 0.839 |
|  | Coastal/Sea | 1.45 (0.79–2.66) | 0.230 |
|  | Mountains/Plateau | 0.81 (0.43–1.52) | 0.511 |
| **Education** | |  |  |
|  | No education | 1.00 (reference) |  |
|  | Primary | 1.26 (0.87–1.81) | 0.220 |
|  | Secondary | 1.11 (0.75–1.65) | 0.605 |
|  | Higher | 2.71 (1.56–4.71) | <0.001 |
| **Marital status** | |  |  |
|  | Never married | 1.00 (reference) |  |
|  | Married/living together | 0.65 (0.39–1.07) | 0.093 |
|  | Formerly married | 0.65 (0.35–1.21) | 0.171 |
| **Currently working** | |  |  |
|  | No | 1.00 (reference) |  |
|  | Yes | 0.90 (0.71–1.16) | 0.425 |
| **Parity** | |  |  |
|  | 0 | 1.00 (reference) |  |
|  | 1–2 | 0.63 (0.39–1.02) | 0.059 |
|  | 3–4 | 0.45 (0.26–0.79) | 0.006 |
|  | ≥5 | 0.65 (0.32–1.32) | 0.232 |
| **Household size** | |  |  |
|  | ≤4 | 1.00 (reference) |  |
|  | 5–6 | 1.22 (0.95–1.56) | 0.121 |
|  | ≥7 | 1.49 (1.10–2.04) | 0.011 |
| **Household motorcycle/scooter ownership** | | |  |
|  | No | 1.00 (reference) |  |
|  | Yes | 0.69 (0.49–0.97) | 0.032 |
| **Household car/truck ownership** | |  |  |
|  | No | 1.00 (reference) |  |
|  | Yes | 0.67 (0.44–1.02) | 0.064 |
| **Drinking-water source type** | |  |  |
|  | Unimproved/other source type | 1.00 (reference) |  |
|  | Improved source type | 0.77 (0.52–1.12) | 0.166 |
| **Alcohol consumption in past month** | |  |  |
|  | No | 1.00 (reference) |  |
|  | Yes | 0.79 (0.55–1.14) | 0.203 |

*Sensitivity analysis restricted to women aged 20–49 years. Adult underweight was defined as BMI <18.5 kg/m². The model used the same complex survey design and final covariate set as the primary analysis. OR, odds ratio; CI, confidence interval; MDD-W, minimum dietary diversity for women.*

**Supplementary Table S6.** Multicollinearity diagnostics for the final multivariable model

| **Model term** | **df** | **GVIF** | **GVIF^(1/(2×df))** |
| --- | --- | --- | --- |
| MDD-W | 1 | 1.10 | 1.05 |
| Household wealth quintile | 4 | 2.65 | 1.13 |
| Age group | 6 | 2.71 | 1.09 |
| Geographical region | 4 | 1.60 | 1.06 |
| Education | 3 | 1.93 | 1.12 |
| Marital status | 2 | 5.03 | 1.50 |
| Currently working | 1 | 1.09 | 1.05 |
| Parity | 3 | 7.61 | 1.40 |
| Household size | 2 | 1.20 | 1.05 |
| Motorcycle/scooter ownership | 1 | 1.13 | 1.06 |
| Car/truck ownership | 1 | 1.33 | 1.16 |
| Drinking-water source type | 1 | 1.20 | 1.10 |
| Alcohol consumption in past month | 1 | 1.02 | 1.01 |

*GVIF, generalized variance inflation factor; df, degrees of freedom. For categorical predictors with more than 1 df, GVIF^(1/(2×df)) provides a dimension-adjusted measure that is comparable across model terms. The largest adjusted GVIF was 1.50, indicating no substantial multicollinearity among variables retained in the final model. Diagnostics were calculated using the complete-case final-model design matrix (n=9,134).*
